# Establishing Minimally Clinically Important Differences for the Orthostatic Hypotension Questionnaire (OHQ)

**DOI:** 10.1101/2025.08.02.25332827

**Authors:** Horacio Kaufmann, Jose-Alberto Palma, Ross Vickery, Lucy Norcliffe-Kaufmann, Beiyao Zheng, David Lewin, Tadhg Guerin

## Abstract

**Purpose:** Establish the minimally clinically important difference (MCID) for the Orthostatic Hypotension Questionnaire (OHQ).

**Background:** Neurogenic orthostatic hypotension (nOH) causes disabling symptoms that impair daily function and quality of life. The OHQ is a validated patient reported outcome with a symptom assessment (OHSA) and daily activity scale (OHDAS), widely used in clinical trials, despite the MCID being unestablished.

**Methods:** We analyzed data from two phase 3, randomized placebo-controlled trials (SEQUOIA and REDWOOD), evaluating ampreloxetine for symptomatic nOH in patients with Parkinson disease, multiple system atrophy, and pure autonomic failure. Using anchor-based and distribution-based methods, we calculated the MCID for the total OHQ score, OHSA and OHDAS composite subscales, and for the single dizziness/lightheadedness question (OHSA1).

**Results:** The analysis included 184 subjects from SEQUOIA and 128 from REDWOOD. The total OHQ MCID for improvement was a reduction of 0.9 to 1.2 points and for worsening was an increase of 0.7 to 1.1 points. The MCID for the OHSA composite ranged from a reduction of 0.9 to 1.3 points for improvement and an increase of 0.7 to 1.1 points for worsening. For the single item OHSA1, the MCID was a reduction of 2.0 to 3.0 points for improvement and an increase of 1.0 point for worsening. Due to poor correlation with the symptom-based anchors, a reliable MCID for the OHDAS component was not established.

**Conclusion:** These MCID thresholds for the OHQ, OHSA and OHSA item 1 alone, enhance the interpretability of scores and support their use in evaluating clinical benefit.

## INTRODUCTION

Neurogenic orthostatic hypotension (nOH) is a chronic, debilitating condition characterized by a drop in blood pressure upon standing, resulting from impaired norepinephrine release from sympathetic nerve terminals [9, 17]. It is commonly associated with neurodegenerative synucleinopathies, such as Parkinson disease (PD), multiple system atrophy (MSA), and pure autonomic failure (PAF), as well as other autonomic neuropathies of autoimmune or genetic causes. Core symptoms result from hypoperfusion of the organs, which can severely compromise daily function and reduce quality of life [9, 10, 13, 15]. Symptom relief remains the primary therapeutic goal in clinical trials and routine care.

The Orthostatic Hypotension Questionnaire (OHQ) was developed as a psychometric tool to capture the patient experience by quantifying symptomatic burden and functional limitations caused by nOH. Over the last decade, it has been widely adopted to assess treatment efficacy in both clinical practice and therapeutic trials [2–8, 11, 14]. The OHQ is currently the only patient-reported outcome (PRO) with regulatory precedent for drug approval in nOH trials [7].

The OHQ prompts the patient to look back over the past week and recall the severity of six core symptoms of nOH and their impact on four functional activities. However, the minimally clinically important difference (MCID), i.e., the smallest change in a score that reflects a meaningful improvement or worsening from the patient’s perspective, had not yet been established for the OHQ. This makes it difficult to predict what constitutes a meaningful therapeutic impact over time. Regulatory guidance emphasizes the importance of assessing meaningful within-patient changes, rather than solely between-group differences, to determine the clinical relevance of a new therapy [1]. Thus, understanding the MCID of the OHQ will bridge an important knowledge gap.

Our objective was to define the MCID thresholds for the OHQ, for both worsening and improvement. The analysis included data from two completed, placebo-controlled phase 3 clinical trials: SEQUOIA and REDWOOD. Both evaluated the efficacy of ampreloxetine (10 mg/day), a long-acting norepinephrine transporter inhibitor, in treating symptomatic nOH [12]. Establishing the MCID will improve the interpretability of OHQ scores and enhance the assessment of treatment efficacy in both clinical trials and practice.

## METHODS

### Study Population

We analyzed pooled data from two completed, sequential, multicenter, phase 3 randomized, placebo-controlled trials. SEQUOIA (NCT03750552) and REDWOOD (NCT03829657). Both trials were designed to evaluate the efficacy of 10 mg once per day oral ampreloxetine in participants with symptomatic nOH. The main entry criteria were: 1) a diagnosis of PD, MSA or PAF; and 2) symptomatic nOH. As part of the entry criteria participants were required to have a minimal symptom severity score of 4 or above on OHQ item 1, the first question that captures symptoms of cerebral hypoperfusion. The diagnosis was confirmed by an Enrollment Steering Committee of 3 independent neurologists. The SEQUOIA study was designed to show benefit and included a run-in 4-week, double-blind, randomized control trial. After completion, participants rolled over into REDWOOD, which was designed to show worsening with a 16-week enriched open-label phase followed by a six-week randomized, double-blind withdrawal phase. The trial designs and analysis plans were developed in consultation with the FDA. Additional participants could enter REDWOOD as new patients. Eligibility criteria were the same in both trials. Participants were randomized to 10 mg/day ampreloxetine vs. matching placebo 1:1 in both blind phases.

### The Orthostatic Hypotension Questionnaire (OHQ)

Both trials with ampreloxetine used the OHQ as the primary endpoint. OHQ scores were collected in a standardized protocol, following a schedule of procedures, which included the participant first watching a video explaining the association between low blood pressure, standing, and symptoms. Scores were collected on a tablet device. The OHQ consisted of ten questions and two components: the 6-item Orthostatic Hypotension Symptom Assessment (OHSA) scale and the 4-item Orthostatic Hypotension Daily Activity Scale (OHDAS). Each item was scored on an 11-point scale from 0 (no symptoms or interference) to 10 (worst possible symptoms or complete interference) [7]. Participants had the option to mark “cannot do for other reasons”. Higher scores indicate worse symptoms.

### OHQ Items and Domains

We set out to analyze the MCID of the OHQ as well as each of its components. The anchoring analysis focused on the OHQ, OHSA and OHDAS composite scores along with OHSA item 1 which has been previously used for drug approval in this therapeutic area. The OHSA and OHDAS composite scores were developed through psychometric testing of construct validity to assess the severity of core symptoms and symptom impact of nOH. The OHSA composite was calculated by averaging the scores of the first 6 questions of the OHQ which assess 1) dizziness/lightheadedness/near-fainting, 2) visual problems, 3) difficulty concentrating, 4) weakness, 5) fatigue, and 6) coat-hanger pain. These symptoms relate to hypoperfusion of the organs, often cited by patients as distressing, and thus considered clinically relevant. The OHDAS composite was calculated by averaging the scores of questions 7 to 10 of the OHQ which assess the impact of these symptoms on: 7) standing for a short time, 8) standing for a long time, 9) walking for a short time, and 10) walking for a long time. The OHQ composite score averages all 10 items. Question 1 on the OHQ (OHSA item 1 “dizziness, lightheadedness, or near fainting”) was analyzed individually as it captures what is thought to be the cardinal symptom of nOH due to cerebral hypoperfusion and was required as a minimal symptom severity score at study entry.

### Anchor-Based Analysis

Anchor-based methods are designed to establish the limit of a clinically meaningful change in a PRO measure. These methods assess the relationship between changes in the target PRO and changes in external “anchor” measures that reflect related clinical concepts. When sufficiently correlated, these anchors help define thresholds for meaningful improvement or worsening from the patient’s experience perspective.

To determine what constitutes a clinically meaningful change in the OHQ scores, we used two patient-reported anchors: the Patient Global Impression of Change (PGI-C) from the SEQUOIA study and the Patient Global Impression of Severity (PGI-S) from the REDWOOD study. These questionnaires were applied at key efficacy assessment visits. PGI-C is a 5-point scale assessing symptom change since starting treatment (as much better, a little better, no change, a little worse, or much worse). The PGI-S is a 5-point scale reflecting symptom severity over the past week (as none, mild, moderate, severe, or very severe).

Data from both placebo and ampreloxetine arms in the randomized blind periods were pooled, regardless of treatment assignment. Prior to conducting anchor-based analyses, we evaluated the relationship between the anchor measures and the relevant OHQ score using Spearman’s correlation coefficients. A correlation of approximately 0.5 or more was required to ensure the anchor was meaningfully associated with the relevant OHQ score [16].

The primary anchor-based analysis identified the median change in the score corresponding to a one-category shift from “no change” on the PGI-C and PGI-S, interpreted as the minimal clinically important difference. We used empirical cumulative distribution functions (eCDFs) to determine the score change at which 50% of patients reached the anchor-defined improvement or worsening threshold. Categories with fewer than fifteen patients were combined or excluded.

In parallel, we used receiver operating characteristic (ROC) curves were generated to determine the score change that best discriminated patients with a ≥1-category improvement or worsening. The optimal threshold was defined as the point minimizing the sum of squared values for 1−sensitivity and 1−specificity.

### Distribution-Based Analysis

To complement the anchor-based approach, we conducted a distribution-based analysis to estimate the minimum detectable change in the scores that can be reliably distinguished from measurement error. For multi-item scales (OHQ composite, OHSA composite, OHDAS composite), this was calculated using the standard error of measurement (SEM), defined by the formula:

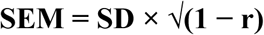

where **SD** is the standard deviation of OHQ, OHSA, or OHSDAS composite scores at baseline, and **r** is the internal consistency reliability, estimated using Cronbach’s alpha for OHSA items 1 through 6 for OHSA and using items 1 through 10 for the OHQ. Because the internal consistency cannot be calculated on a single-item measure, the half-standard deviation (0.5 * SD) method was used for OHSA item 1. These calculations were performed separately at baseline in the SEQUOIA trial and at the randomized withdrawal baseline in the REDWOOD trial. Only MCID thresholds that exceeded the relevant measurement error estimate (SEM or 0.5 * SD) were considered clinically interpretable.

### Statistical Analyses

All statistical analyses were performed using SAS version 9.4 and SAS/STAT version 15.1 (SAS Institute, Cary, NC, USA). The primary anchor-based analysis calculated the median change in the OHQ scores across PGI-C and PGI-S categories, with additional analyses using ROC curves and eCDFs as supportive methods.

### Ethical Considerations

Both trials were conducted in accordance with the Declaration of Helsinki and received ethical approval from the appropriate institutional review boards (IRB). All participants provided written informed consent, including consent for future research use of their data.

All participants provided informed consent for using their data in future research on nOH and related conditions, ensuring ethical compliance in data utilization. Both the SEQUOIA and REDWOOD studies were conducted under the oversight of IRBs, and all data were anonymized to protect participant confidentiality.

## RESULTS

### Patient Characteristics

A total of 184 patients from SEQUOIA and 128 from REDWOOD were included in the anchoring analysis. All patients met enrollment criteria for PD, MSA, or PAF and nOH. The protocol pre-specified a minimum enrolment of at least 40% MSA patients. The mean age of the population was 67 years, with a slight male predominance (see table 1 for baseline demographic and clinical characteristics).

**Table 1.**
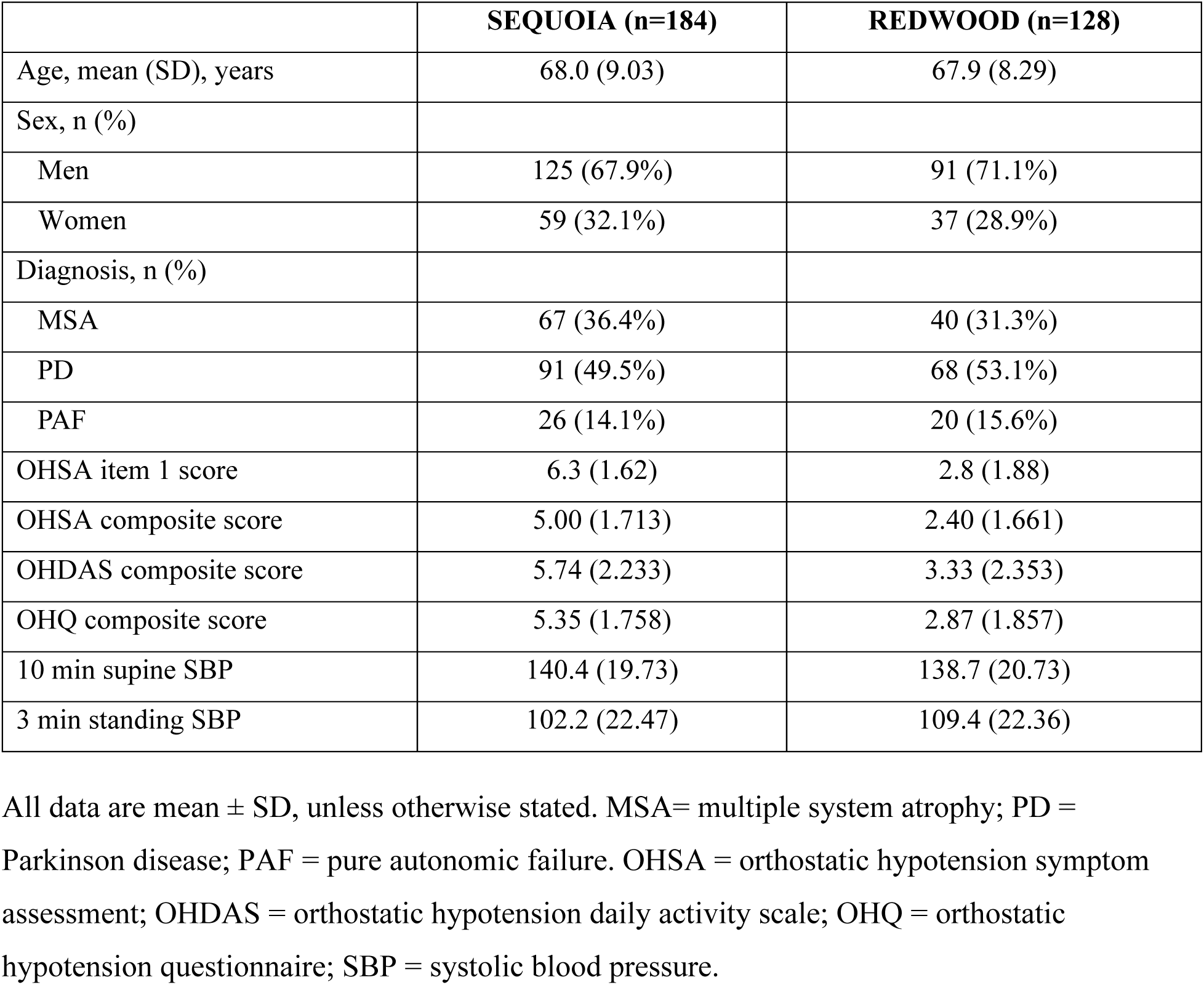
Baseline Characteristics of Study Participants with nOH at baseline.

**Table 2:** Median change in OHQ Scores by PGI-C categories at Week 4 (SEQUOIA Study)

| **PGI-C Category** | **n** | **Median Change in OHQ Composite Score** | **Median Change in OHSA Composite Score** | **Median Change in OHSA item1** |
| --- | --- | --- | --- | --- |
| Much better | 36 | -3.0 | -2.2 | -3.0 |
| A little better | 57 | -1.4 | -1.9 | -3.0 |
| No change | 51 | -0.5 | -0.6 | 0.0 |
| A little worse | 18 | 0.2 | 0.5 | 1.0 |
| Much worse | 14 | 1.0 | 0.9 | 1.0 |

PGI-C = patient global impression of change; OHQ = orthostatic hypotension questionnaire; OHSA = orthostatic hypotension symptom assessment.

**Table 3.** Median change in OHQ Scores by PGI-S categories at Week 6 of Randomized-Withdrawal (REDWOOD Study.

| **PGI-S Category** | **n** | **Median Change in OHQ Composite Score** | **Median Change in OHSA Composite Score** | **Median Change in OHSA item1** |
| --- | --- | --- | --- | --- |
| 3-category improvement | 1 | -0.2 | -1.60 | -1.0 |
| 2-category improvement | 1 | -0.4 | -0.30 | -2.0 |
| 1-category improvement | 13 | -0.1 | -0.20 | -1.0 |
| No change | 58 | -0.15 | 0.00 | 0.0 |
| 1-category worsening | 34 | 0.9 | 0.70 | 1.0 |
| 2-category worsening | 6 | 1.65 | 1.50 | 4.5 |
| 3-category worsening | 1 | 6.3 | 5.50 | 6.0 |

PGI-Severity = patient global impression of severity; OHQ = orthostatic hypotension questionnaire; OHSA = orthostatic hypotension symptom assessment.

### Anchor Viability

To assess the suitability of the PGI-C and PGI-S as external anchors, we evaluated their correlation with changes in OHQ scores across both trials. In the SEQUOIA study, which utilized the PGI-C, Spearman correlation coefficients indicated a strong association with the OHQ composite score (r = 0.64), the OHSA composite score (r = 0.60), and OHSA item 1 (r = 0.61). In the REDWOOD study, which employed the PGI-S, correlations were moderately strong for the OHSA composite score (r = 0.50), OHSA item 1 (r = 0.55), and somewhat lower for the OHQ composite score (r = 0.45). These results supported the use of these anchors for estimating MCIDs for the OHQ and OHSA composite scores along with OHSA item 1, as they met or exceeded the threshold of r ≈ 0.5 recommended for anchor-based analyses.

In contrast, the anchors were deemed unsuitable for deriving a reliable MCID for the OHDAS composite score. In SEQUOIA, patients who rated themselves as “a little worse” on the PGI-C often showed contradictory improvement in OHDAS scores. Similarly, in REDWOOD, the correlation between change in OHDAS scores and PGI-S was weak (r = 0.33). Moreover, patients reporting improvement on the PGI-S unexpectedly demonstrated greater worsening on the OHDAS score compared to those reporting no change. These inconsistencies suggest a conceptual mismatch between the symptom-based anchors and the activity-based OHDAS scale, which likely reflects limitations in using symptom severity anchors to assess functional impairment.

### Identified Anchor Thresholds

The following MCIDs were identified for the OHQ composite score, the OHSA composite score and OHSA item 1. There were insufficient numbers of patients improving on the PGI-S in REDWOOD to provide an estimate of minimal improvement in that study.

- **OHQ Composite Score:** In SEQUOIA, the threshold was −0.9 for minimal improvement and +0.7 for minimal worsening. In REDWOOD, the threshold for minimal worsening was +1.1.
- **OHSA Composite Score:** In the SEQUOIA study, a change of −1.3 points (’a little better’ vs. ‘no change’) was the threshold for minimal improvement, while +1.1 points (‘a little worse’ vs. ‘no change’) indicated minimal worsening. In the REDWOOD study, a change of +0.7 points represented minimal worsening.
- **OHSA Item 1:** In SEQUOIA, the threshold was −3.0 for minimal improvement and +1.0 for minimal worsening. In REDWOOD, the threshold for minimal improvement and worsening were −1.0 and +1.0 respectively.

### Distribution-Based Analysis

The estimated measurement error was used to ensure anchor-derived thresholds were credible. In SEQUOIA / REDWOOD, the SEM was 0.69 / 0.58 for the OHQ composite and 0.85 / 0.65 for the OHSA composite. For the single-item OHSA1, the half standard deviation was 0.81 / 0.94. All MCID thresholds proposed above exceeded these measurement error values.

### ROC Curve Analysis

In the SEQUOIA dataset, ROC curve analysis identified a change of −1.2 for the OHQ composite score (AUC = 0.824), a change of −0.9 points on the OHSA composite score (AUC = 0.826; Figure 3), and a change of −2.0 for OHSA1 (AUC = 0.826) as the optimal thresholds for detecting a clinically meaningful improvement (≥1-category change on the PGI-C). From the REDWOOD dataset, the optimal worsening threshold was 0.8 for the OHQ composite score (AUC=0.778) and 1.0 for OHSA1 (AUC=0.787). A worsening threshold for OHSA composite from REDWOOD fell below the SEM and was excluded.

**FIGURE 1:**
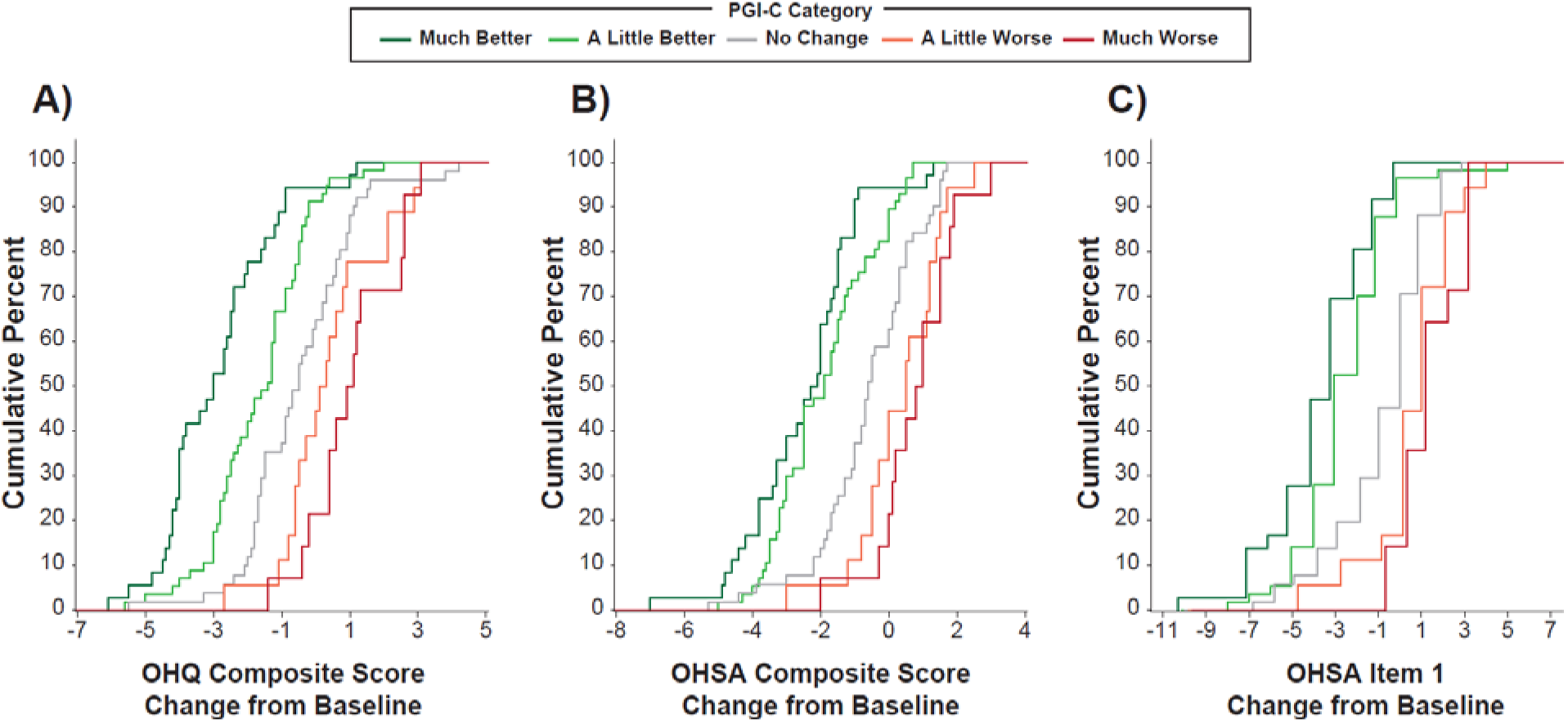
Anchor-based analysis of scores at Week 4 in SEQUOIA study. A) Shows change in OHQ total scores according to PGI-Change answers. B) Shows change in OHSA-composite symptom burden scores derived from the first 6 symptom-based items according to PGI-Change answers. C) Shows in change in Item 1 scores for dizziness/light-headedness alone according to PGI-Change answers. OHQ = Orthostatic Hypotension Questionnaire; PGI-C = patient global impression of change; OHSA = Orthostatic hypotension symptom assessment.

**FIGURE 2:**
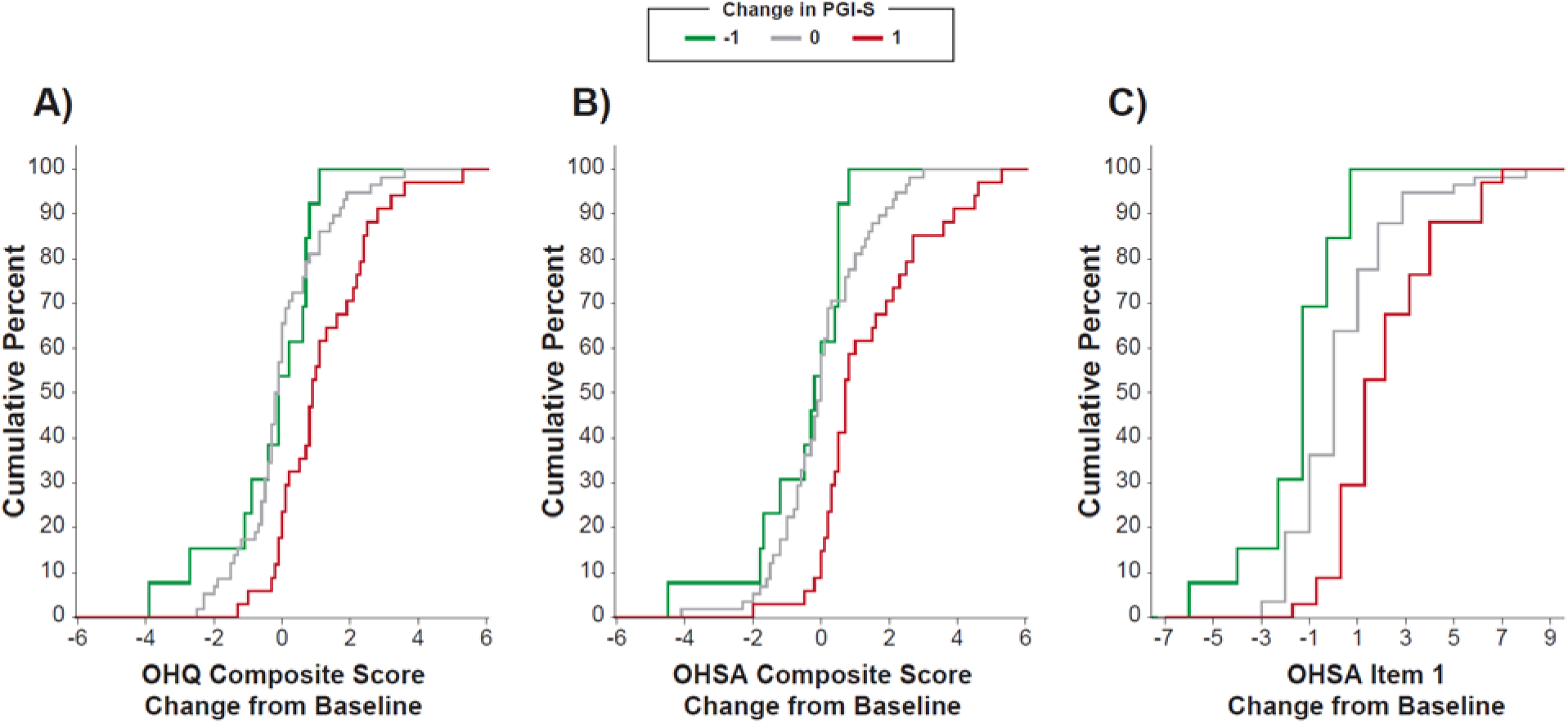
Anchor-based analysis of Scores at week 6 in REDWOOD study. A) Shows change in OHQ total scores according to PGI-Severity answers. B) Shows change in OHSA-composite symptom burden scores derived from the first 6 symptom-based items according to PGI-Severity answers. C) Shows in change in Item 1 scores for dizziness/light-headedness alone according to PGI-Severity answers. Note, only changes in PGI-S of −1, 0, and 1 are shown due to insufficient data in other categories to combine groups to a sample size of ≥ 15 patients. OHQ = Orthostatic Hypotension Questionnaire; PGI-S = patient global impression of severity; OHSA = Orthostatic hypotension symptom assessment.

**FIGURE 3:**
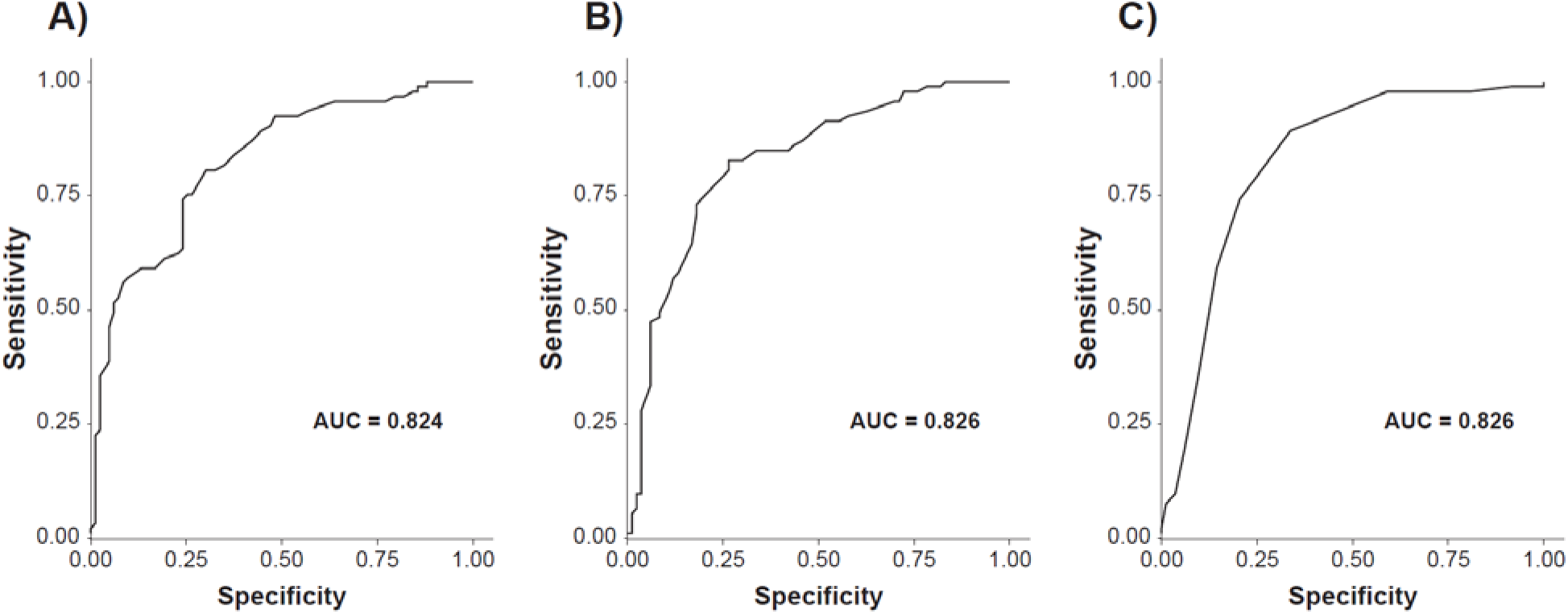
Sensitivity and specificity of changes in OHQ Scores in the detection of symptom improvement at week 4 in SEQUOIA study. A) Shows receiver operating characteristic (ROC) curve for sensitivity and specificity to detect change in OHQ total scores. Note, a change of −1.2 in the total OHQ score (i.e., symptomatic improvement) had the best-combined sensitivity and specificity (AUC=0.824) to detect a change equal to or greater than 1 category in the PGI-C. B) Shows ROC curve with the sensitivity and specificity of a change in OHSA Composite Score to detect an improvement in the PGI-C. A change of −0.9 in the OHSA Composite Score (i.e., symptomatic improvement) had the best-combined sensitivity and specificity (AUC=0.826) to detect a change equal to or greater than 1 category in the PGI-C. C) shows ROC curve for item 1, dizziness/light-headedness alone, where a change of −2.0 had the best-combined sensitivity and specificity (AUC=0.826) to detect a change equal to or greater than 1 category in the PGI-C. AUC = area under the curve; OHSA = Orthostatic Hypotension Symptom Assessment; PGI-C = Patient Global Impression of Change.

### Summary of MCID Thresholds

Table 4 presents a summary of the derived thresholds from both studies. The final MCID range for improvement on the OHQ composite score was −0.9 to −1.2 points, and for worsening, +0.7 to +1.1 points. For the OHSA composite score, the MCID range for improvement was −0.9 to −1.3 points, and for worsening, +0.7 to +1.1 points. For OHSA1, the MCID was −2.0 to −3.0 for improvement and +1.0 for worsening.

**Table 4.** Derived thresholds are indicative of minimally clinically important changes in nOH patients.

| OHQ Score | Method | Improvement threshold | Worsening threshold |
| --- | --- | --- | --- |
| OHSA Composite | PGI-C from SEQUOIA Study | -1.3 | 1.1 |
|  | PGI-S from REDWOOD Study | N/A | 0.7 |
|  | ROC from SEQUOIA Study | -0.9 | N/A |
|  | **Identified MCID** | **-0.9 to -1.3** | **0.7 to 1.1** |
| OHQ Composite | PGI-C from SEQUOIA Study | -0.9 | 0.7 |
|  | PGI-S from REDWOOD Study | N/A | 1.1 |
|  | ROC from SEQUOIA/REDWOOD Studies | -1.2 | 0.8 |
|  | **Identified MCID** | **-0.9 to -1.2** | **0.7 to 1.1** |
| OHSA1 | PGI-C from SEQUOIA Study | -3.0 | 1.0 |
|  | PGI-S from REDWOOD Study | N/A | 1.0 |
|  | ROC from SEQUOIA/REDWOOD Studies | -2.0 | 1.0 |
|  | **Identified MCID** | **-2.0 to -3.0** | **1.0** |

OHQ = orthostatic hypotension questionnaire; PGI-Severity = patient global impression of severity; PGI-C = patient global impression of change; OHSA = orthostatic hypotension symptom assessment; ROC = receiver operating curve; MCID = minimal clinical important difference.

## DISCUSSION

This study established the MCID for the OH-Questionnaire (OHQ); specifically – for the total score (OHQ composite), symptom domain component (OHSA composite), and Item 1 (dizziness, lightheadedness). This analysis provides the thresholds for detecting benefit and worsening using data from two large placebo-controlled phase 3 trials in nOH. For the OHQ composite score, a 0.9 to 1.2-point reduction represents clinically important improvement, while an increase of 0.7 to 1.1 points reflects meaningful worsening. Similarly, for the OHSA composite score, a decrease of 0.9 to 1.3 points reflects a clinically important improvement, whereas an increase of 0.7 to 1.1 points indicates worsening. For Item 1 (OHSA#1, dizziness/lightheadedness), a 2 to 3-point reduction represents meaningful improvement, while a 1-point increase indicates meaningful worsening. These thresholds were derived and validated using both anchor-based and distribution-based methodologies, supporting their robustness and clinical relevance.

The identification of these MCID thresholds represents a critical advance in the interpretation of patient-reported outcomes in nOH. By establishing what constitutes meaningful symptomatic improvement or deterioration, patients are better equipped to report, evaluate treatment efficacy to clinicians and researchers and guide their care.

Our study also highlighted the challenge in defining an MCID for the activity based OHDAS component using a symptom anchor. This is likely because they address different components – symptoms and activities, which correlate closely with disease stage. The PGI-C and PGI-S anchors ask about the change in overall symptoms, whereas the OHDAS measures the impact of symptoms on daily activities, which represents a conceptual mismatch between the anchor and the outcome. This mismatch between symptom-based anchors and the activity focus of the OHDAS led to inconsistent findings in our anchor analysis. From a clinical perspective, standing and walking may also be influenced by factors unrelated to orthostatic hypotension, due to motor deficits, concomitant medications, and motor fluctuations. These factors confound the activity-based questions measure most in patients with PD and MSA, due to the motor decline.

A major strength of the study is the use of data acquired from two complementary trial designs, the SEQUOIA which investigated symptom *benefit* and REDWOOD which investigated symptom *worsening*. This design is particularly well suited for evaluating symptomatic benefit over time especially when using a randomized withdrawal approach as the standard for long-term assessments. This dual trial approach offered a more comprehensive basis for MCID estimation. The rigorous screening process for patient selection included a tilt-table test confirmation of a >20/10 systolic/diastolic blood pressure fall at 3 minutes upright, a minimal symptom cut off (Item 1 >4 points). The clinical diagnosis was carefully reviewed with a written narrative to support consensus criteria adjudicated by an independent enrollment steering committee. This approach, combined with careful site selection, oversight, ongoing investigator training, and continuous data monitoring, provides a robust analysis of an international multi-center dataset (76 clinical sites in 19 countries). The MICD threshold ranges identified through this analysis has direct applicability to future therapeutic trials in this neurotherapeutic space for autonomic disorders.

This study has some limitations. Our analyses were based on data from two clinical trials that investigated a single agent, ampreloxetine. Although these cohorts were representative of typical nOH populations, further validation of these MCID thresholds in other trial settings and real-world populations is needed.

Moreover, although the anchor instruments (PGI-C and PGI-S) ask about symptom changes, they rely on patients’ impressions and are inherently subjective and rely on recall. Future studies should explore the use of additional or more objective anchors—such as activity monitoring or physiologic measures—to further validate changes. The enrollment criteria restricted patients with seated plasma norepinephrine levels <100 pg/ml and required a minimum symptom severity score at baseline. It is not known if these ranges are generalizable to patients with more severe peripheral autonomic nerve degeneration or milder nOH symptoms scores.

The range in MCID thresholds underscores the difficulty in assessing symptoms of nOH in the patient population, which show diurnal variations, and only a modest correlation between symptom severity and blood pressure acquired at a single office visit. Nevertheless, prioritizing symptoms over orthostatic blood pressure values aligns with the regulatory standards and treatment practice. The OHSA composite scale allows the patient to score their global symptom burden rather than a single-item approach used in prior trials. This likely better reflects the patient experience as it allows participants to derive a score from a constellation of symptoms rather than report on only one symptom as bothersome like dizziness/lightheadedness alone (Item 1).

Our analysis provides validated MCID thresholds that serve as a valuable tool for interpreting patient-reported outcomes, both in clinical practice and in the regulatory trials designed to evaluate new therapies. Detecting a meaningful difference in symptom benefit and worsening will enhance decision-making and inform the design and interpretation of clinical trials targeting symptomatic relief in nOH. This study approach lays a foundation for standardizing clinical trial endpoints by facilitating interpretation, to help with regulatory review of emerging treatments in autonomic medicine. Ongoing analysis is needed to confirm the applicability of these thresholds as ways to optimize patient-centered care for patients living with nOH.

## Data Availability

All data produced in the present study are available upon reasonable request to the authors

## Funding

This study was funded by Theravance Biopharma.

## Financial Interests

H. Kaufmann is Editor-in-Chief of Clinical Autonomic Research, published by Springer-Nature, and serves as PI of studies sponsored by Biogen MA Inc. (TRACK MSA, S19-01846) and Vaxxinity Inc. (UB-312, S22-01332). He has received consultant fees from Takeda Pharmaceutical Company Ltd, Ono Pharma UK Ltd, Theravance Biopharma US LLC., and Parexel. He receives royalties from Up to Date. He receives research support from NIH-NINDS, the Food and Drug Administration, the Familial Dysautonomia Foundation, and the HSAN IV Foundation. J.-A. Palma reports research funding from the NIH. R. Vickery is an employee of Theravance Biopharma Ireland Limited and is a shareholder. L. Norcliffe-Kaufmann and B. Zheng are employees of Theravance Biopharma US, LLC and are shareholders. D. Lewin is a contractor for Theravance Biopharma Ireland Limited. T. Guerin is an employee of Theravance Biopharma Ireland Limited and is a shareholder.

## Author contributions

HK, JAP, RV, LNK, & TG conceived and designed the research; BZ, DL, & TG analyzed data; HK, JAP, LNK, & TG drafted, edited & revised the manuscript; HK, RV, LNK & TG approved the final version of the manuscript.

## Notes

### Clinical Trial

NCT03750552; NCT03829657

### Author Declarations

Ethics committee/IRB of Advarra (IRB Registration #: 00000971) gave ethical approval for this work

